# Non-pharmaceutical interventions to reduce influenza transmission in households: a systematic review and meta-analysis

**DOI:** 10.1101/2024.09.10.24313390

**Authors:** Jessica Y. Wong, Wey Wen Lim, Justin K. Cheung, Caitriona Murphy, Eunice Y. C. Shiu, Jingyi Xiao, Dongxuan Chen, Yanmin Xie, Mingwei Li, Hualei Xin, Michelle Szeto, Sammi Choi, Benjamin J. Cowling

**Affiliations:** World Health Organization Collaborating Centre for Infectious Disease Epidemiology and Control, School of Public Health, Li Ka Shing Faculty of Medicine, The University of Hong Kong, Hong Kong Special Administrative Region, China; Laboratory of Data Discovery for Health Limited, Hong Kong Science and Technology Park, New Territories, Hong Kong Special Administrative Region, China

**Author notes:** **Corresponding author:** Prof. Benjamin J. Cowling, School of Public Health, Li Ka Shing Faculty of Medicine, The University of Hong Kong, 7 Sassoon Road, Pokfulam, Hong Kong.

**Keywords:** Influenza, Non-pharmaceutical interventions, Households

## Abstract

**Background:** Influenza pandemic plans often recommend non-pharmaceutical interventions (NPIs) in household settings, including hand hygiene and face masks. We reviewed the evidence supporting the recommendations of these measures to prevent the spread of influenza in households.

**Methods:** We performed systematic reviews between 26 May and 30 August 2022 in Medline, PubMed, EMBASE, and CENTRAL to identify evidence for the effectiveness of selected measures recommended by representative national influenza pandemic plans. We prioritized evidence from randomized controlled trials. Fixed-effects models were used to estimate the overall effects. Systematic reviews were registered in the OSF registry (https://osf.io/8kyth).

**Results:** We selected 9 NPIs for evidence review. We identified 9 randomized-controlled trials related to hand hygiene and face masks in household settings. 2 studies reported that measures could delay the introduction of influenza virus infections into households. However, we did not identify evidence from randomized controlled trials that indicated a substantial effect of hand hygiene and face masks in preventing the spread of pandemic influenza within households.

**Conclusion:** Limited evidence indicated that within-household measures may likely be effective only when implemented before or as soon as possible after symptom onset in an infected case. Improving the evidence base for NPIs in households and elsewhere is a continuing priority.

**Funding:** World Health Organization and the Strategic Topic Grants Scheme

## INTRODUCTION

The threat posed by the next influenza A pandemic has not diminished in the wake of the COVID-19 pandemic. It is important to adapt influenza pandemic plans in light of experiences from the COVID-19 pandemic. Given the delays in the availability of specific vaccines and limited stockpiles of influenza antivirals in many locations, non-pharmaceutical interventions (NPIs) – also referred to as public health and social measures – will continue to provide the first line of defense in the next influenza pandemic, just as they did at the start of the COVID-19 pandemic [1].

Influenza virus infections spread mainly through inhalation of infectious respiratory particles that can occur during close contact between individuals, and one of the settings responsible for a considerable fraction of all influenza transmission is households [2]. In the 2009 influenza A(H1N1)pdm09 pandemic, some studies estimated that around one-third of all transmission events occurred in households [3]. NPIs in households could, therefore, make a major contribution to containment or mitigation efforts. We reviewed the scientific evidence supporting NPIs that might be recommended to reduce influenza transmission in households.

## METHODS

### Selection of NPIs

We reviewed the websites of national public health organizations from around the world to determine which NPIs might be recommended in households during influenza epidemics or pandemics (Table S1). Two to three countries were selected as a sample from each continent to capture snapshots of country-specific recommendations for NPIs to mitigate the spread of influenza in households. From this, we identified a list of NPIs that could be assessed in evidence reviews.

### Search strategy

We then conducted systematic reviews between 26 May and 30 August 2022 to evaluate the effectiveness of these selected measures on influenza virus transmission in the household setting. These systematic reviews followed the Preferred Reporting Items for Systematic Reviews and Meta-Analyses (PRISMA) guidelines. The protocol was registered in the Open Science Framework (OSF) registry under the registration number https://osf.io/8kyth. Four databases (Medline, PubMed, EMBASE, and CENTRAL) were searched for literature in all languages with specific search terms (Table S2).

### Study selection

For each review, two authors screened titles of all papers identified by the search strategy independently. Abstracts for potentially relevant papers and the full texts of manuscripts were assessed for eligibility. We aimed to identify studies of the efficacy of each measure against laboratory-confirmed influenza outcomes in “private” household settings, and defined a private household (denoted as “household” hereafter) as two or more individuals living, not necessarily related, under the same unit with common housekeeping (i.e. providing food for themselves) [4]. We prioritized evidence from randomized controlled trials (RCTs) as they provide the highest quality of evidence. For measures with a lack of RCTs with laboratory-confirmed influenza outcomes, we also searched for observational studies on laboratory-confirmed influenza, influenza-like illness (ILI), and respiratory illness outcomes (such as acute respiratory illness or ARI). If a published systematic review was identified through our search, we updated the review using pre-defined search terms and evaluated literature published after the search date of the previous review. Because the relative importance of modes of influenza transmission might vary in different household settings, studies that were conducted in “institutional” households (such as dormitories for students and homes for the elderly) whose need for shelter and subsistence is being provided by a common authority were excluded.

### Statistical analysis

Meta-analyses were performed for interventions with a sufficient number of studies. The efficacy or effectiveness of measures in preventing laboratory-confirmed influenza was measured by risk ratios (RRs). Overall effects were estimated in pooled analyses with fixed-effects models. No overall effect was generated if there was considerable heterogeneity based on an estimated *I*^2^ statistic ≥75%. The Appendix includes additional details of the search strategies (Tables S1 and S2), selection of articles (Figures S1-S9), and summaries of the selected articles (Tables S3 and S4).

## RESULTS

### National public health guidance on NPIs in households

We reviewed the websites of national public health organizations from 15 countries, specifically: Ghana, Nigeria and South Africa in Africa; China, Singapore and South Korea in Asia; Germany, Italy and United Kingdom in Europe; Canada and United States in North America; Australia and New Zealand in Oceania; and Peru and Brazil in South America (Table 1). NPIs that were implemented could be broadly categorized as personal protective measures, environmental measures or other measures which included measures such as hand hygiene, surface disinfection or physical distancing respectively. For personal protective measures, all selected countries except Germany recommended hand hygiene and respiratory etiquette in household settings, while around half of the countries (e.g., China, South Korea and Italy) recommended the use of face masks. None of the sampled countries recommended face shields. Similarly, around half of the countries (e.g., South Africa, China and Germany) recommended surface and object cleaning or ventilation or both as environmental measures in household settings, and none recommended humidification. Finally, all countries recommended the isolation of sick individuals and physical distancing in household settings during influenza epidemics or pandemics.

**Table 1:** Recommendations of household-related non-pharmaceutical interventions in different countries.

| Continent | Country | Personal protective measures |  |  |  | Environmental measures |  |  | Other measures |  |
| --- | --- | --- | --- | --- | --- | --- | --- | --- | --- | --- |
|  |  | Hand hygiene | Respiratory etiquette | Face masks | Face shields | Surface and object cleaning | Ventilation | Humidification | Isolation of sick individuals | Physical distancing |
| Africa | Ghana | ✓ | ✓ |  |  |  |  |  | ✓ | ✓ |
|  | Nigeria | ✓ | ✓ |  |  |  |  |  | ✓ | ✓ |
|  | South Africa | ✓ | ✓ | ✓ |  | ✓ |  |  | ✓ | ✓ |
| Asia | China | ✓ | ✓ | ✓ |  | ✓ | ✓ |  | ✓ | ✓ |
|  | Singapore | ✓ | ✓ |  |  |  |  |  | ✓ | ✓ |
|  | South Korea | ✓ | ✓ | ✓ |  |  |  |  | ✓ | ✓ |
| Europe | Germany |  |  | ✓ |  |  | ✓ |  | ✓ | ✓ |
|  | Italy | ✓ | ✓ | ✓ |  |  |  |  | ✓ | ✓ |
|  | United Kingdom | ✓ | ✓ |  |  |  |  |  | ✓ | ✓ |
| North America | Canada | ✓ | ✓ |  |  | ✓ |  |  | ✓ | ✓ |
|  | United States | ✓ | ✓ | ✓ |  | ✓ | ✓ |  | ✓ | ✓ |
| Oceania | Australia | ✓ | ✓ |  |  | ✓ |  |  | ✓ | ✓ |
|  | New Zealand | ✓ | ✓ |  |  |  | ✓ |  | ✓ | ✓ |
| South America | Brazil | ✓ | ✓ | ✓ |  |  |  |  | ✓ | ✓ |
|  | Peru | ✓ | ✓ | ✓ |  | ✓ |  |  | ✓ | ✓ |

Country-specific recommendations on NPIs during influenza epidemics or pandemics were generally disseminated through national health agency websites in the form of general health information or formal guidelines for influenza (Table S1) [5, 6]. Recommendations in four countries were updated after the A(H1N1)pdm09 pandemic [6–8], while recommendations for the other 11 countries were updated during the COVID-19 pandemic (Table S1) [9, 10].

### Systematic review of intervention studies

From the review of national recommendations, we constructed a list of 9 NPIs including those that have been recommended and some that have not (Table 2). We identified a total of 23,001 articles for title and abstract screening across the 9 NPIs and 800 full-text articles were retrieved and reviewed (Figures S1–S9). For hand hygiene, 576 articles were reviewed, 62 full-text articles were screened, and 7 intervention studies were identified for the meta-analysis. For face masks, 1,890 articles were reviewed, 151 full-text articles were screened, and 7 intervention studies were identified for the meta-analysis. No intervention studies were identified for the other 7 NPIs. After removing duplicates for studies based on hand hygiene and face masks, 9 unique intervention studies were included in the review (Tables 2, S3–S4).

**Table 2:**
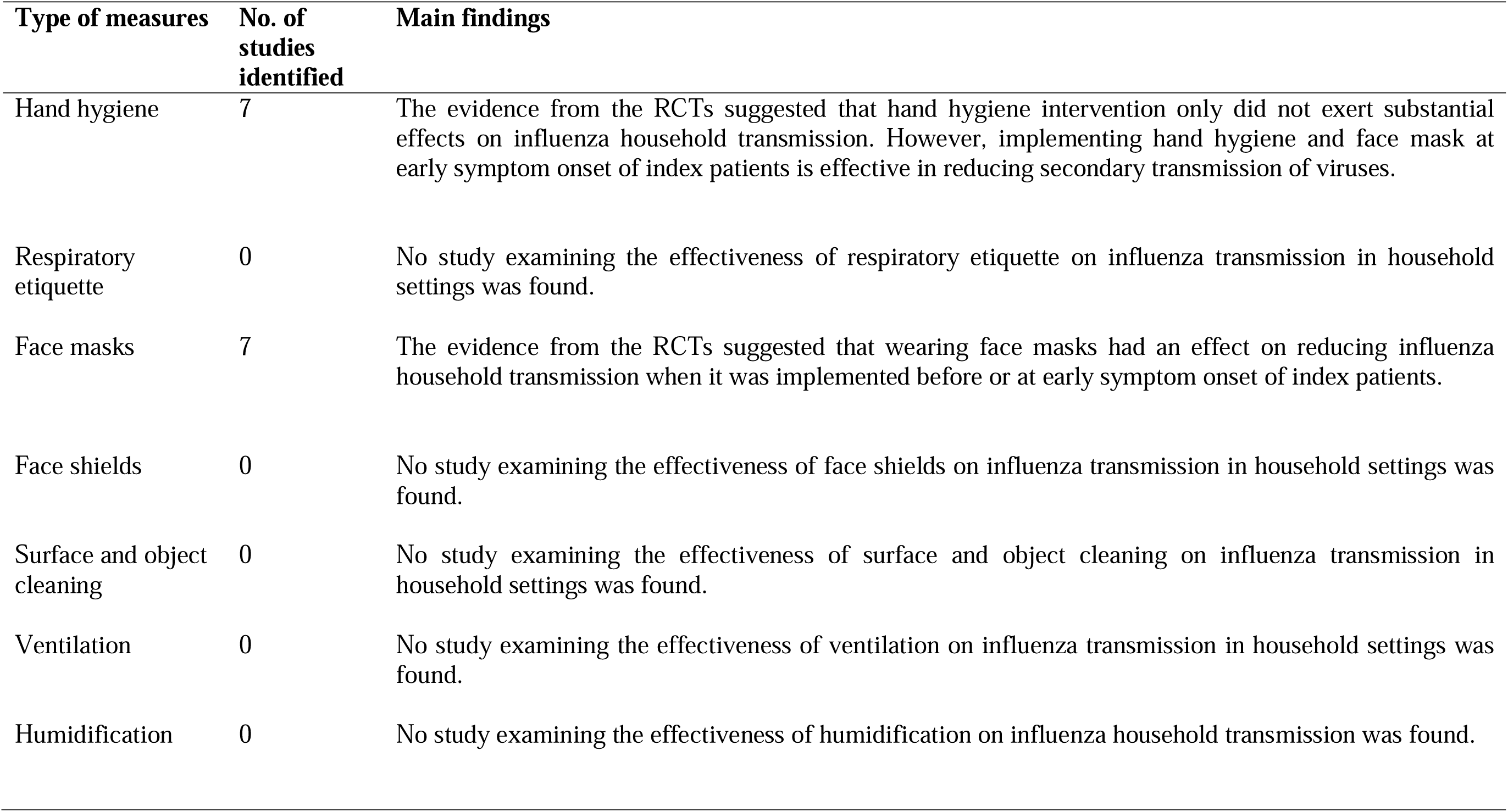
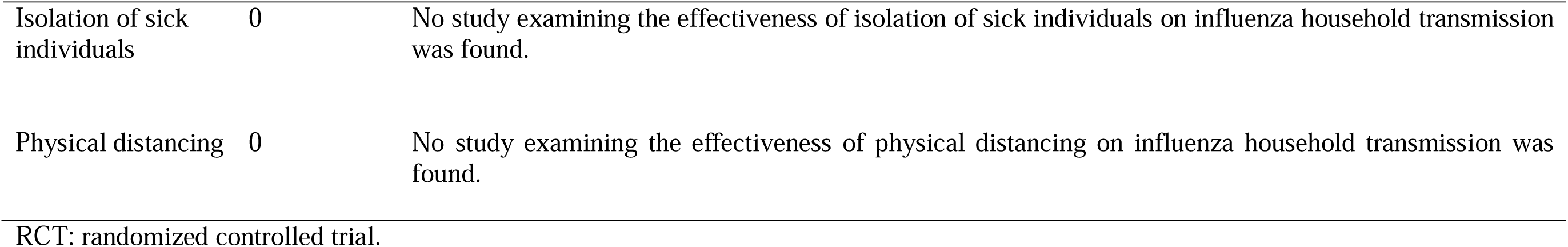
Summary of literature searches for systematic review on non-pharmaceutical interventions in household settings for influenza.

| **Type of measures** | **No. of studies identified** | **Main findings** |
| --- | --- | --- |
| Hand hygiene | 7 | The evidence from the RCTs suggested that hand hygiene intervention only did not exert substantial effects on influenza household transmission. However, implementing hand hygiene and face mask at early symptom onset of index patients is effective in reducing secondary transmission of viruses. |
| Respiratory etiquette | 0 | No study examining the effectiveness of respiratory etiquette on influenza transmission in household settings was found. |
| Face masks | 7 | The evidence from the RCTs suggested that wearing face masks had an effect on reducing influenza household transmission when it was implemented before or at early symptom onset of index patients. |
| Face shields | 0 | No study examining the effectiveness of face shields on influenza transmission in household settings was found. |
| Surface and object cleaning | 0 | No study examining the effectiveness of surface and object cleaning on influenza transmission in household settings was found. |
| Ventilation | 0 | No study examining the effectiveness of ventilation on influenza transmission in household settings was found. |
| Humidification | 0 | No study examining the effectiveness of humidification on influenza household transmission was found. |
| Isolation of sick individuals | 0 | No study examining the effectiveness of isolation of sick individuals on influenza household transmission was found. |
| Physical distancing | 0 | No study examining the effectiveness of physical distancing on influenza household transmission was found. |

RCT: randomized controlled trial.

### Personal protective measures: hand hygiene, respiratory etiquette, face masks, and face shields

We identified seven RCTs, six of which were included in the meta-analysis, to assess the efficacy of hand hygiene against transmission of laboratory-confirmed influenza in household settings with at least one case, with 5,118 participants (Figure 1; Tables S3) [11–13]. The study by Levy et al [12] was excluded in the meta-analysis because the number of secondary influenza virus infections were reported in terms of number of households instead of number of individuals. An overall pooled effect of hand hygiene only, hand hygiene combined with face masks, and hand hygiene with or without face masks was estimated. Results from our meta-analysis on RCTs did not provide evidence to support a protective effect of hand hygiene only against transmission of laboratory-confirmed influenza (RR: 1.07; 95% CI: 0.85-1.35; p-value: 0.58; *I*^2^ =48%). Although the pooled analysis did not identify a significant effect of hand hygiene on laboratory-confirmed influenza overall, some household transmission studies reported that initiating hand hygiene intervention earlier after symptom onset in the index case might be more effective in preventing secondary cases in the household settings [11, 13].

**Figure 1:**
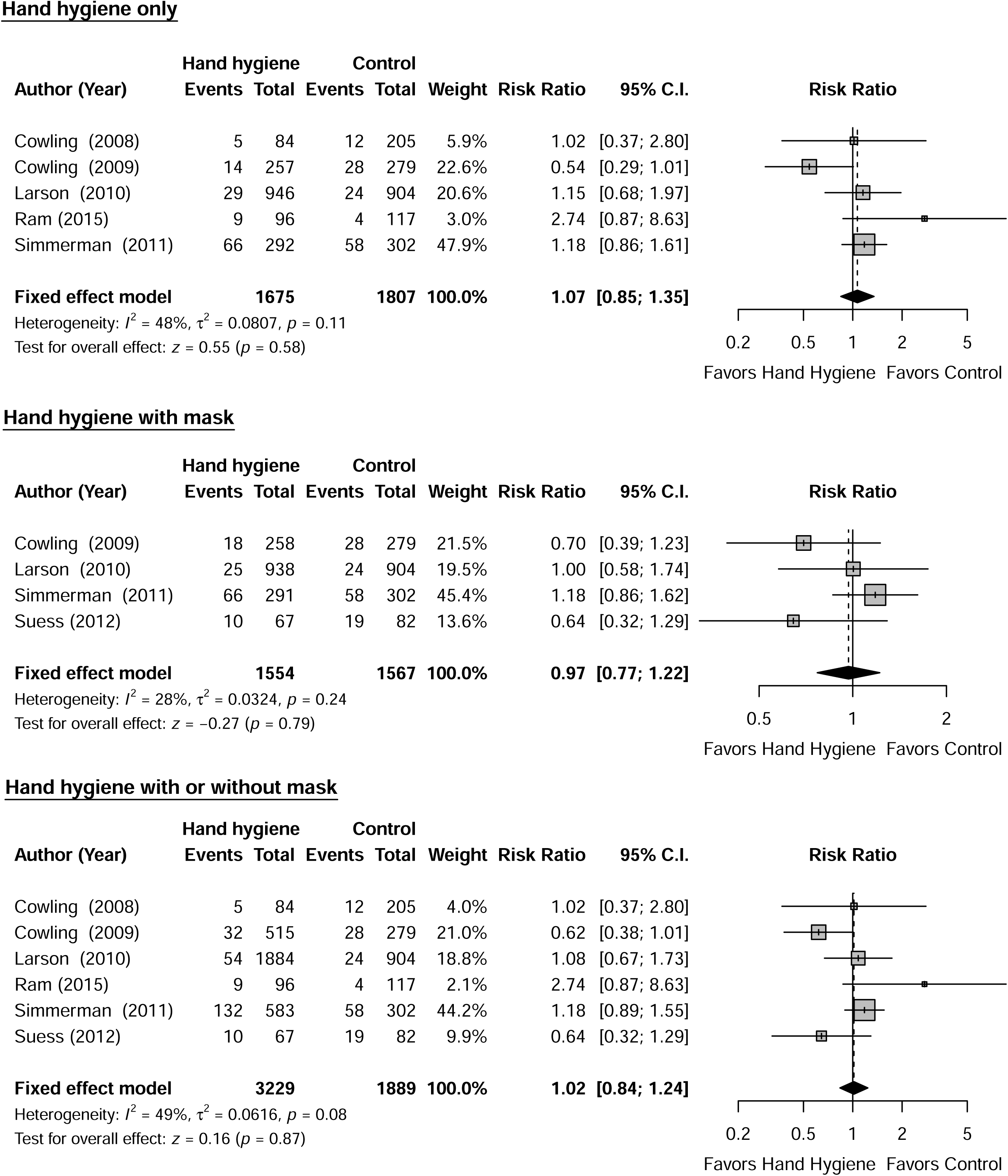
Meta-analysis of risk ratios for the effect of hand hygiene with or without face mask use on laboratory-confirmed influenza from 6 randomized controlled trials with 5,118 participants. (A) Hand hygiene alone; (B) Hand hygiene and face mask; (C) Hand hygiene with or without face mask. Pooled estimates were not generated if there was high heterogeneity (*I^2^* >75%). Squares indicate risk ratio for each of the included studies, horizontal line indicates 95% CIs, dashed vertical line indicates pooled estimation of risk ratio, and diamond indicates pooled estimation of risk ratio. Diamond width corresponds to the 95% CI. The study by Levy et al was excluded in the meta-analysis but included in the review as its number of secondary infections are measured in households instead of participants [12].

In our systematic review, we identified seven RCTs that reported estimates of the effectiveness of face masks in reducing laboratory-confirmed influenza virus infections in household settings (Table S4) [11, 13]. Five of these trials investigated the masking of all household members, regardless of symptom presentation, and we were therefore unable to distinguish the potential effects of face masks worn by infected vs uninfected individuals [11, 13]. Despite results not being statistically significant, a trial on face masks reported a lower risk of ILI and laboratory-confirmed influenza infection among those with medical mask use, and similar results were reported in an earlier study. In the pooled analysis, there was no statistically significant reduction in influenza transmission with the use of face masks only (RR: 0.59; 95% CI: 0.32-1.10; p-value 0.10; *I*^2^ =16%) (Figure 2). Study designs in the seven household studies were slightly different: one trial provided face masks and P2 respirators for household members only, another trial evaluated the use of face masks as source control for infected individuals only, and the remaining five trials provided face masks for the infected individuals as well as their household members (Table S4) [11, 13]. Only two household studies reported a statistically significant reduction in secondary laboratory-confirmed influenza virus infections, when face masks were worn within 36 hours of symptom onset [11, 13]. Most household studies were underpowered due to small sample sizes, and some studies reported suboptimal adherence in the face mask group.

**Figure 2:**
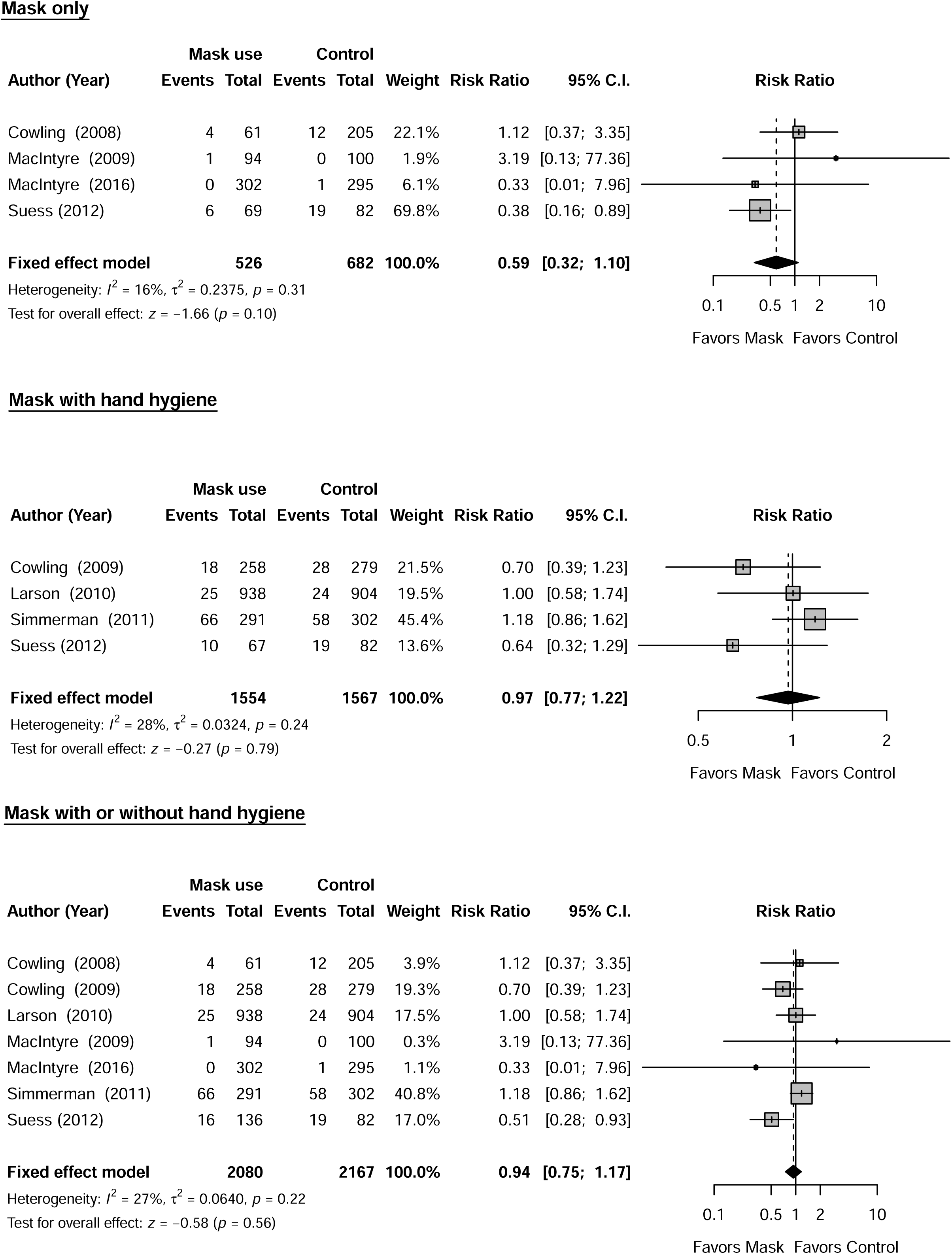
Meta-analysis of risk ratios for the effect of face mask use with or without hand hygiene on laboratory-confirmed influenza from 7 randomized controlled trials with 4,247 participants. (A) Face mask use alone; (B) Face mask and hygiene; (C) Face mask with or without hand hygiene. Pooled estimates were not generated if there was high heterogeneity (*I^2^*>75%). Squares indicate risk ratio for each of the included studies, horizontal line indicates 95% CIs, dashed vertical line indicates pooled estimation of risk ratio, and diamond indicates pooled estimation of risk ratio. Diamond width corresponds to the 95% CI.

We did not identify any published intervention studies on the effectiveness of respiratory etiquette and face shields in reducing the risk of laboratory-confirmed influenza in household settings.

### Environmental measures: surface and object cleaning, ventilation, humidification

We did not identify any published intervention studies that quantified the effectiveness of modifying humidity, surface and object cleaning, or ventilation in reducing influenza transmission in household settings.

### Isolation of sick individuals and physical distancing

We did not identify any published intervention studies on the effectiveness of isolation policies for sick individuals and physical distancing measures in reducing the risk of laboratory-confirmed influenza in household settings.

## DISCUSSION

Prevention and control of respiratory virus infections in households is an important yet relatively underexplored area of research. Guidelines for infection prevention and control of seasonal and pandemic influenza in healthcare settings are well established [14]. During the COVID-19 pandemic, several guidelines on infection control and prevention in households using NPIs were issued by health authorities alongside guidance for self-care and family care. For example, the World Health Organization Q&A webpage on “Home care for families and caregivers” recommends donning medical masks while sharing a space with someone with COVID-19, staying at least 1 meter away from the sick person, and opening windows to bring fresh air into the sick person’s room where possible [15]. Although the feasibility of these measures may depend on living conditions, forward planning for the possibility of having a household member who is sick with an infectious disease is prudent even in inter-pandemic periods [16].

Among household settings, hand hygiene, face masks, respiratory etiquette, surface and object cleaning and ventilation are feasible NPIs to implement during an influenza epidemic or pandemic. With hand hygiene and face masks as recommended hygiene practices to limit the spread of respiratory virus infections within the household, the effectiveness of such measures could be enhanced through public health campaigns that boost compliance [17]. Similarly, respiratory etiquette should be highly feasible in household settings, and an improvement in compliance has been demonstrated among school children after piloting an educational intervention in one study in elementary schools [18]. It should also be feasible to implement surface and object cleaning in the household due to the low cost of implementation and accessibility of common household cleaning agents. Given the potential for aerosol transmission of respiratory viruses including influenza [19], improving ventilation should be considered except perhaps for households in areas with poor outdoor air quality or when this would substantially increase heating costs. When household members are sick, it should often be feasible to isolate those sick individuals and increase physical distancing, for example by avoiding spending time in the same rooms or eating separately with them [20], although it may be more challenging in households with crowded living conditions.

In this review, we did not find evidence to support a substantial protective effect of personal protective measures, environmental measures, isolation of sick individuals or physical distancing measures in reducing influenza transmission in household settings. Although these measures have mechanistic plausibility of reducing influenza transmission based on our knowledge of how influenza is transmitted between individuals [21, 22], randomized trials of hand hygiene and face marks in household settings have not demonstrated protection against laboratory-confirmed influenza. There were no RCTs on respiratory etiquette, face shields, modifying humidity, ventilation, isolation policies for sick individuals and physical distancing in household settings.

Despite a lack of intervention studies on measures other than face masks and hand hygiene, we identified an observational study on the association between indoor humidity and influenza transmission, suggesting a potential role of humidification in controlling transmission of influenza [23] although there are also potential harms of humidification which would need to be considered, such as increasing mold. Other studies suggested that surface and object cleaning using common household agents, indoor ventilation and voluntary self-isolation were effective measures in reducing influenza transmission by inactivating influenza viruses in the environment or decreasing the transmission risk [24]. Another retrospective cohort study found that daily use of chlorine or ethanol-based disinfectant was effective (OR: 0.23, 95% CI: 0.07, 0.84) in reducing COVID-19 household transmission, and similarly for face mask use (OR: 0.21; 95% CI: 0.06, 0.79) and surface disinfection when the measures were implemented before symptom onset of the primary case [20]. The disinfection of surfaces also has an established impact on prevention of other infectious diseases such as gastrointestinal diseases [25].

When devising strategies to reduce influenza transmission in households, it is important to understand the basic transmission dynamics of influenza virus infections. In the next pandemic, important information on transmission dynamics of the novel strain could be provided by timely First Few Hundred studies [26] and household transmission studies [27]. If the transmission dynamics of the new pandemic strain are similar to that of H1N1pdm09 and current interpandemic strains, we can note the following four properties. First, infectiousness is thought to peak at around the same time as when symptoms appear [28]. Second, infectiousness likely declines rapidly within a few days after peak based on viral culture data [29] despite viral RNA continuing to be detectable by PCR typically for more than a week [30]. Third, only a fraction of influenza virus infections result in fever, and while fever and cough may be a relatively more specific syndrome for influenza, it is not particularly sensitive in the general community as contrasted with its higher sensitivity in individuals who seek medical attention with respiratory symptoms [31]. Fourth, the role of asymptomatic and pre-symptomatic transmission has been controversial but recent reports from South Africa [32] and Hong Kong [33] indicate that these may comprise a substantial fraction of all influenza transmission, with asymptomatic and pre-symptomatic transmission also playing an important role in COVID-19 transmission [34]. This fundamental knowledge of infectiousness profiles would imply that early intervention is essential to reduce transmission, and early intervention should not be limited to individuals with a fever and cough but could be triggered by other less specific symptoms. Rapid antigen tests done in the household could help to distinguish influenza from other viral infections and might even be considered for use in exposed individuals to identify influenza virus infection before any symptoms appear.

There are a number of limitations to our review. First, in our analysis of the effectiveness of face masks and hand hygiene we did not review observational data as a higher level of evidence from randomized controlled trials were available. Other studies have reviewed observational data and concluded that these two measures likely have small to moderate effects on transmission [35]. Second, we focused on measures to prevent the spread of influenza within the household in this review. There is limited evidence on the degree of reductions in transmission in households when personal protective measures (e.g., wearing face masks plus frequent hand hygiene) are used in combination with other measures like isolation of sick household members. The effectiveness of different cleaning products at different concentrations in deactivating or eliminating influenza virus in household environments remains unclear. Third, increased influenza activity is associated with cold temperatures, low indoor humidity and rainy seasons [36]. Further investigation could clarify the effectiveness of NPIs by different seasonal patterns (such as indoor crowding during colder months). Finally, we observed low to moderate levels of heterogeneity in our meta-analyses of hand hygiene and face masks (Figures 1-2). We could determine whether these differences were artefactual or real, perhaps related to differences in the adherence of measures in various populations or the time delay between symptom onset of an infected case and the implementation of a measure [37]. Further work could attempt to identify additional factors that explain this heterogeneity, for example, by exploring very different estimates of effectiveness of measures based on the same population during similar time periods, or conducting subgroup analyses by the time delay between symptom onset and measure implementation.

Improved evidence is needed on all of the measures included in our review. Given the effect sizes in our meta-analysis of hand hygiene and face masks (Figures 1 and 2), any future RCTs of these interventions in households would likely need to be very large to be adequately powered to detect a relative reduction in the risk of infection of approximately 10% [38]. To avoid contamination of interventions, cluster randomized trials, in which each household is randomized to receive either the intervention or control, could be used to assess the effect of the intervention in reducing the transmission of influenza in households [11]. A promising area for randomized trials or cluster-randomized trials in the household setting is the effect of physical distancing on influenza transmission, either by alternating within-home isolation strategies or comparing the feasibility and effectiveness of physical distancing in housing areas with different population densities. Surveys about the feasibility of each measure in local contexts are also important to inform national-level recommendations on home care and/or voluntary self-isolation or quarantine [39].

In conclusion, although our study found limited evidence to support a substantial protective effect of personal protective measures, environmental measures, isolation of sick persons or physical distancing measures in controlling influenza transmission in the household setting, these measures have mechanistic plausibility based on our knowledge of person-to-person transmission of influenza [21, 22]. Future investigations on transmission dynamics of influenza would be helpful in preparing guidelines and evidence-based recommendations for household transmission in the next pandemic. Although our review focused on NPIs to be used during influenza pandemics, these results could also be applicable to intense seasonal influenza epidemics.

## Data Availability

All data produced in the present work are contained in the manuscript

## ACKNOWLEDGMENTS

The authors thank Julie Au for administrative support.

## DATA SHARING STATEMENT

The data that support the findings of this study are available upon request.

## CONTRIBUTIONS

All authors meet the ICMJE criteria for authorship. The study was conceived by BJC, and JYW. JYW and JKC analyzed the data. JYW wrote the first draft of the manuscript. All authors provided critical review and revision of the text and approved the final version.

