## Supplementary Materials for "Non-pharmaceutical interventions to reduce influenza transmission in households: a systematic review and meta-analysis"

SUPPLEMENTARY MATERIAL

This supplementary material provides additional information on the data and search strategies used.

### **Table S1. List of websites of national public health organizations**

| **Continent** | **Country** | **Public health organization** | **Reference** |
| --- | --- | --- | --- |
| Africa | Ghana | Ghana Food and Drugs Authority | ^1^ |
|  | Nigeria | Nigeria Centre for Disease Control and Prevention | ^2^ |
|  | South Africa | The National Institute for Communicable Diseases | ^3^ |
| Asia | China | Chinese Center for Disease Control and Prevention | ^4^ |
|  | Singapore | Ministry of Health | ^5^ |
|  | South Korea | Korea Disease Control and Prevention Agency | ^6^ |
| Europe | Germany | The Robert Koch Institute | ^7^ |
|  | Italy | Ministry of Health | ^8^ |
|  | United Kingdom | National Health Service | ^9^ |
| North America | Canada | Public Health Agency of Canada | ^10^ |
|  | United States | Centers for Disease Control and Prevention | ^11^^,^ ^12^ |
| Oceania | Australia | Communicable Diseases Network Australia | ^13^ |
|  | New Zealand | Health New Zealand \| Te Whatu Ora | ^14^ |
| South America | Brazil | Ministry of Health | ^15^ |
|  | Peru | Ministry of Health | ^16^ |

### **Table S2. Search strategies for systematic reviews**

| **Intervention** | **Search strategy** | **Search terms** | **Search date** |
| --- | --- | --- | --- |
| Hand hygiene | We initially conducted a review of the 12 articles included in Xiao ^17^ studying the effect of hand hygiene interventions on prevention of laboratory-confirmed influenza in household settings. We then conducted an updated search in PubMed, MEDLINE, EMBASE and CENTRAL up to 26 May 2022, regardless of language. Literature in languages other than English were excluded during full-text screening and only articles with randomized controlled trials (RCTs) study design and examined the effect of hand hygiene interventions on laboratory-confirmed influenza in household settings were included. Systematic reviews and meta-analyses, as well as studies involving non-household settings were excluded. Two reviewers (JX and ES) independently screened the titles, abstracts and full texts to identify articles for inclusion. After data extraction, quality assessment of the evidence was carried out based on the GRADE framework. | #1: "hand hygiene" OR "hand washing" OR "handwashing" OR "hand-washing" OR "hand-wash" OR "hand wash" OR "handwash" OR "hand sanitize" OR "hand sanitizers" OR "hand sanitizer" OR "hand rub" OR "handrub" OR "hand rubbing" OR "hand cleansing" OR "hand cleans" OR "hand cleanser" OR "hand disinfectant" OR "hand disinfectants" OR "hand disinfection" OR "hand soap" OR "hand wipe"  #2: "influenza" OR "flu" OR “SARS CoV-2” OR “COVID-19” OR “COVID” OR “respiratory syncytial virus” OR “RSV” OR “parainfluenza” OR “adenovirus” OR “rhinovirus” OR “respiratory virus” OR “respiratory infection” OR “respiratory tract infection” OR “respiratory illness”  #3: #1 AND #2 | 26 May 2022 |
| Respiratory etiquette | We conducted a systematic review on the effect of respiratory etiquette in preventing respiratory virus infections in household settings. A literature search was then conducted on 29 June 2022 in 4 databases: PubMed, MEDLINE, EMBASE and CENTRAL to identify any potential RCTs. No language limit was applied for the literature search. RCTs and other types of epidemiological studies were included if they evaluated the effectiveness of respiratory etiquette in reducing the risk of laboratory-confirmed respiratory viruses in household settings, Two reviewers independently (ES and ZX) screened the titles, abstracts, and full texts to identify articles for inclusion. | #1: "respiratory hygiene" OR "cough etiquette" OR "respiratory etiquette"  #2: "influenza" OR "flu" OR "SARS-CoV-2" OR "COVID-19" OR "COVID" OR "respiratory syncytial virus" OR "RSV" OR "parainfluenza" OR "adenovirus" OR "rhinovirus" OR "respiratory virus" OR "respiratory infection" OR "respiratory tract infection" OR "respiratory illness"  #3: #1 AND #2 | 29 June 2022 |
| Face masks | On 28 May 2022, a literature search was conducted in 4 databases: PubMed, MEDLINE, EMBASE and CENTRAL to identify any potential RCTs. No language limit was applied for the literature search. Reference lists of the retrieved articles were also reviewed for additional articles for this review. Only RCTs were included, and the included trials must evaluate the effectiveness of face mask wearing (with or without the combination of other inventions as one intervention) on the mitigation of respiratory viruses in household settings using the incidence of laboratory-confirmed influenza or other respiratory virus infections as the study outcome. Two reviewers independently (ES and ZX) screened the titles, abstracts, and full texts to identify articles for inclusion. | #1: "facemask" OR "facemasks" OR "mask" OR "masks" OR "respirator" OR "respirators"  #2: "influenza" OR "flu" OR “SARS CoV-2” OR “COVID-19” OR “COVID” OR “Respiratory syncytial virus” OR “RSV” OR “parainfluenza” OR “adenovirus” OR “rhinovirus” OR “respiratory virus” OR “respiratory infection” OR “respiratory tract infection” OR “respiratory illness”  #3: #1 AND #2 | 28 May 2022 |
| Face shields | We conducted a systematic review on the effect of face shields in preventing respiratory virus infections in household settings. A literature search was conducted on 7 August 2022 in 4 databases: PubMed, MEDLINE, EMBASE and CENTRAL to identify any potential studies. No language limit was applied for the literature search. RCTs and other types of epidemiological studies were included if they evaluated the effectiveness of face shields on reducing secondary transmission in household settings. Two reviewers independently (ES and ZX) screened the titles, abstracts, and full texts to identify articles for inclusion. | #1: "face shield" OR "face shields"  #2: "influenza" OR "flu" OR "SARS-CoV-2" OR "COVID-19" OR "COVID" OR "respiratory syncytial virus" OR "RSV" OR "parainfluenza" OR "adenovirus" OR "rhinovirus" OR "respiratory virus" OR "respiratory infection" OR "respiratory tract infection" OR "respiratory illness"  #3: #1 AND #2 | 7 August 2022 |
| Surface and object cleaning | We conducted a systematic review on the effect of surface and object cleaning in preventing respiratory virus infections in household settings. PubMed, Medline, EMBASE and CENTRAL were searched for articles on 4 July 2022.  Study selection criteria were studies reporting the effect of surface and object cleaning intervention with no intervention in preventing respiratory virus infections in household settings. RCTs and other types of epidemiological studies were included if they aimed to study the effect of surface and object cleaning on laboratory-confirmed influenza, ILI or other respiratory illness. Simulation studies, recommendations, and commentaries or editorials were excluded. Articles describing any surface and object cleaning related interventions were included. No language limits were applied.  Two independent reviewers (ES and ZX) reviewed titles, abstracts and full texts. After confirmed included studies, data extraction and evidence quality assessment were performed by the reviewers. The GRADE framework was used to evaluate the effect of surface and object cleaning on influenza. | #1: "surface" OR "surfaces" OR "object" OR "objects" OR "fomite" OR "fomites" OR "environment" OR "environmental"  #2: "clean" OR "cleans" OR "cleaning" OR "cleanse" OR "cleansing" OR "disinfect" OR "disinfects" OR "disinfection" OR "disinfecting" OR "wipe" OR "wipes" OR "sanitize" OR "sanitizes" OR "sanitizing" OR "sanitation" OR "sterilize" OR "sterilizes" OR "sterilizing" OR "sterilization" OR "sterilise" OR "sterilises" OR "sterilising" OR "sterilisation" OR "decontaminate" OR "decontaminates" OR "decontaminating" OR "decontamination"  #3: "influenza" OR "flu" OR "SARS-CoV-2" OR "COVID-19" OR "COVID" OR "respiratory syncytial virus" OR "RSV" OR "parainfluenza" OR "adenovirus" OR "rhinovirus" OR "respiratory virus" OR "respiratory infection" OR "respiratory tract infection" OR "respiratory illness"  #4: #1 AND #2 AND #3 | 4 July 2022 |
| Ventilation | We conducted a systematic review on the effect of ventilation in preventing respiratory virus infections in household settings. A literature search was conducted on 2 August 2022 in 4 databases: PubMed, MEDLINE, EMBASE and CENTRAL. No language limit was applied for the literature search. RCTs and other types of epidemiological studies were included if they evaluated the effectiveness of ventilation on mitigating laboratory-confirmed influenza respiratory viruses in household settings. Two reviewers independently (ES and ZX) screened the titles, abstracts, and full texts to identify articles for inclusion. | #1: "ventilation" OR "environment"  #2: "indoor" OR "room"  #3: "influenza" OR "flu" OR "SARS-CoV-2" OR "COVID-19" OR "COVID" OR "respiratory syncytial virus" OR "RSV" OR "parainfluenza" OR "adenovirus" OR "rhinovirus" OR "respiratory virus" OR "respiratory infection" OR "respiratory tract infection" OR "respiratory illness"  #4: #1 AND #2 AND #3 | 2 August 2022 |
| Humidification | We conducted a systematic review on the effect of humidification in preventing respiratory virus infections in household settings. A literature search was conducted on 7 August 2022 in 4 databases: PubMed, MEDLINE, EMBASE and CENTRAL. No language limit was applied for the literature search. RCTs and other types of epidemiological studies were included if they evaluated the effectiveness of humidification on reducing laboratory-confirmed influenza respiratory transmissions in household settings. Two reviewers independently (ES and ZX) screened the titles, abstracts, and full texts to identify articles for inclusion. | #1: "humidity" OR "temperature" OR "environment"  #2: "indoor" OR "room"  #3: "influenza" OR "flu" OR "SARS-CoV-2" OR "COVID-19" OR "COVID" OR "respiratory syncytial virus" OR "RSV" OR "parainfluenza" OR "adenovirus" OR "rhinovirus" OR "respiratory virus" OR "respiratory infection" OR "respiratory tract infection" OR "respiratory illness"  #4: #1 AND #2 AND #3 | 7 August 2022 |
| Isolation of infected individuals | A literature search was conducted using PubMed, MEDLINE, EMBASE and CENTRAL up to 26 May 2022. No language limit was applied for the literature search; however, literature in languages other than English were excluded during full-text screening. The inclusion criteria were studies reporting the effectiveness of isolation on the control of influenza and other respiratory viruses in household settings. No limitation on study design was applied for study inclusion because preliminary works have identified no RCTs for this topic. Systematic reviews and meta-analyses, as well as studies involving clinical settings were excluded. Two reviewers independently (YX and DX) screened the titles, abstracts, and full texts to identify articles for inclusion. | #1 "patient isolation" OR "case isolation" OR "voluntary isolation" OR "home isolation" OR "social isolation" OR "self-isolation"  #2: "influenza" OR "flu" OR “SARS CoV-2” OR “COVID-19” OR “COVID” OR “Respiratory syncytial virus” OR “RSV” OR “parainfluenza” OR “adenovirus” OR “rhinovirus” OR “respiratory virus” OR “respiratory infection” OR “respiratory tract infection” OR “respiratory illness”  #3: #1 AND #2 | 26 May 2022 |
| Physical distancing | A systematic review was conducted using PubMed, MEDLINE, EMBASE and CENTRAL up to 30 August 2022. Studies reporting the effectiveness of physical distancing on the control of influenza or other respiratory viruses in non-health care settings were included. No limitation on language or study design was applied for inclusion as preliminary works identified no RCT for this topic. Systematic review and meta-analyses, as well as studies involving clinical settings, were excluded. Two reviewers (CM and ML) independently screened the titles, abstracts, and full texts to identify articles for inclusion. Quality assessment of evidence was conducted for epidemiological studies. | #1: "physical"  #2: "distancing" OR "distance"    #3: "influenza" OR "flu" OR "SARS-CoV-2" OR "COVID-19" OR "COVID" OR "respiratory syncytial virus" OR "RSV" OR "parainfluenza" OR "adenovirus" OR "rhinovirus" OR "respiratory virus" OR "respiratory infection" OR "respiratory tract infection" OR "respiratory illness"  #4: #1 AND #2 AND #3 | 30 August 2022 |

### **Figure S1. Flowchart of literature search and article selection for hand hygiene**

### **Figure S2. Flowchart of literature search and article selection for respiratory etiquette**

### **Figure S3. Flowchart of literature search and article selection for face masks**

### **Figure S4. Flowchart of literature search and article selection for face shields**

### **Figure S5. Flowchart of literature search and article selection for surface and object cleaning**

### **Figure S6. Flowchart of literature search and article selection for ventilation**

### **Figure S7. Flowchart of literature search and article selection for humidification**

### **Figure S8. Flowchart of literature search and article selection for isolation of sick individuals**

### **Figure S9. Flowchart of literature search and article selection for physical distancing**

### **Table S3. Summary of studies included in the review of hand hygiene**

| **Study** | **Study design** | **Study Period** | **Population and setting** | **Location** | **Transmission mode** | **Intervention** | **Outcome and finding** |
| --- | --- | --- | --- | --- | --- | --- | --- |
| Cowling (2008)^18^ | Cluster-RCT | Feb 2007 to  Sep 2007 | 198 laboratory-confirmed influenza cases and their household contacts recruited from outpatient clinics | Hong Kong | Secondary | Hand sanitizer and education; face mask and education; control: receive same education, but no additional intervention | No significant difference between intervention and control group in laboratory-confirmed influenza and clinical secondary attack rate |
| Cowling (2009)^19^ | Cluster-RCT | Jan 2008 to  Sep 2008 | 407 laboratory-confirmed influenza cases recruited from outpatient clinics, 259 households which included 794 household contacts were further analysed | Hong Kong | Secondary | Hand sanitizer and education; hand sanitizer, face mask and education; control: receive same education, but no additional intervention | Hand hygiene and face mask prevent influenza transmission, but not statistically significant; the interventions significantly reduce influenza transmission within 36 hours of index case symptom onset |
| Larson (2010)^20^ | Cluster- RCT | Nov 2006 to Jun 2008 | 617 households recruited, 509 households were further analysed | New York, USA | Primary and secondary | Hand sanitizer and education; hand sanitizer, face mask and education; control: receive same education, but no additional intervention | No detectable benefit of hand hygiene, face mask and education intervention on influenza prevention. |
| Levy  (2014)^21^ | Cluster-RCT | Jun 2010 to Nov 2010 | 191 households with index children recruited from a public paediatric hospital | Bangkok, Thailand | Secondary | Hand washing; hand washing and face masks: control: no intervention | Less secondary influenza infections in households in the intervention group than control group, but not statistically significant; handwashing reduces surface influenza RNA contamination |
| Ram  (2015)^22^ | Cluster-RCT | Jan 2009 to  Dec 2010 | 384 households with index case- patients recruited from a hospital, among them, 60 index cases were laboratory-confirmed influenza infection | Kishoregoni, Bangladesh | Secondary | Soap and daily handwashing promotion; control: no intervention | Handwashing promotion did not prevent secondary influenza infection in household setting |
| Simmerman (2011)^23^ | Cluster-RCT | Apr 2008 to Aug 2009 | 465 households recruited from a public pediatric hospital, 442 households were further analyzed | Bangkok, Thailand | Secondary | Hand washing and education; hand washing, face masks and education; control: education that was unrelated to personal protective measures, no additional interventions | No significant reduction in rate of secondary influenza infection in control, hand, mask + hand group |
| Suess  (2012)^24^ | Cluster-RCT | Nov 2009 to Jan 2010;  Jan 2011to  Apr 2011 | 84 laboratory-confirmed influenza cases and 218 household contacts recruited by general practitioners or paediatricians | Berlin, Germany | Secondary | Face masks and education; Hand sanitizer, face masks and education; control: education on infection prevention, no additional interventions | Face masks+ hand hygiene could reduce influenza transmission in household setting, result of mask group alone is significant, result of mask+ hand hygiene is not statistically significant |

### **Table S4. Summary of studies included in the review of face masks**

| **Study** | | **Study design** | **Study Period** | **Population and setting** | **Location** | **Transmission mode** | **Intervention** | **Outcome and finding** |
| --- | --- | --- | --- | --- | --- | --- | --- | --- |
| Cowling (2008)^18^ | | Cluster-RCT | Feb to  Sep 2007 | 198 laboratory-confirmed influenza cases and their household contacts recruited from outpatient clinics | Hong Kong, China | Secondary | Face mask and education; hand sanitizer and education; control: receive same education, but no additional intervention | No significant difference between intervention and control group in laboratory-confirmed influenza and clinical secondary attack rate |
| Cowling (2009)^19^ | | Cluster-RCT | Jan to  Sep 2008 | 407 laboratory-confirmed influenza cases recruited from outpatient clinics, 259 households which included 794 household contacts were further analysed | Hong Kong, China | Secondary | Face mask, hand sanitizer and education; Hand sanitizer and education; control: receive same education, but no additional intervention | Face masks and hand hygiene prevent influenza transmission, but not statistically significant; the interventions significantly reduce influenza transmission within 36 hours of index case symptom onset |
| Larson (2010)^20^ | | Cluster- RCT | Nov 2006 to Jun 2008 | 617 households recruited, 509 households were further analysed | New York, USA | Primary and secondary | Hand sanitizer and education; Face mask, hand sanitizer and education; control: receive same education, but no additional intervention | No detectable benefit of face masks, hand hygiene and education intervention on influenza prevention. |
| MacIntyre (2009)^25^ | | Cluster-RCT | Aug 2006 to Oct 2006,  Jun 2007 to  Oct 2007 | 145 laboratory-confirmed influenza case and their adult household contacts | Victoria, Australia | Secondary | Surgical masks; P2 masks; control: no masks used, but no additional interventions | No significant difference in rate of laboratory-confirmed influenza in control, face mask or P2 mask group |
| MacIntyre (2016)^26^ | | Cluster-RCT | Nov 2013 to Jan 2014 | 245 ILI index case and 597 household contacts | Beijing, China | Secondary | Masks; control: no masks, but not additional interventions | Clinical respiratory illness, ILI and laboratory-confirmed viral infections were lower in the mask arm compared to control, but the results were not statistically significant |
| Simmerman (2011)^23^ | | Cluster-RCT | Apr 2008 to Aug 2009 | 465 households recruited from a public pediatric hospital, 442 households were further analyzed | Bangkok, Thailand | Secondary | Face masks, handwashing and education; hand washing and education; control: education that was unrelated to personal protective measures, no additional interventions | No significant reduction in rate of secondary influenza infection in mask+ hand, hand, and control group |
| Suess  (2012)^24^ | | Cluster-RCT | Nov 2009 to Jan 2010;  Jan 2011 to  Apr 2011 | 84 laboratory-confirmed influenza cases and 218 household contacts recruited by general practitioners or paediatricians | Berlin, Germany | Secondary | Face masks and education; Hand sanitizer, face masks and education; control: education on infection prevention, no additional interventions | Face masks+ hand hygiene could reduce influenza transmission in household setting, result of mask group alone is significant, result of mask+ hand hygiene is not statistically significant |

### **Table S5. Grade quality assessment and evidence profile for hand hygiene**

| **Quality assessment** | | | | | | | **No. of patients** | | **Effect** |  |  |
| --- | --- | --- | --- | --- | --- | --- | --- | --- | --- | --- | --- |
| No. of studies | Design | Risk of bias | Inconsistency | Indirectness | Imprecision | Other considerations | Hand hygiene | Control | Risk ratio | Quality | Importance |
| **Effect of hand hygiene intervention on prevention of laboratory-confirmed influenza** | | | | | | |  |  |  |  |  |
| 6 | Randomized controlled trial^1,2^ | No serious risk of bias^3-7^ | Not serious^8^ | No serious indirectness^9^ | Serious^10^ | None | 242/3229 | 145/1889 | 1.02  (0.84, 1.24) | Moderate | Important |

| ^1^ | All studies were randomized controlled trials. |
| --- | --- |
| ^2^ | All studies were cluster-RCTs |
| ^3^ | Four studies reported blinding of study staff including clinical staff, laboratory staff or recruiting physicians. Subjects of all studies were not blinded due to the nature of the study design. |
| ^4^ | Three studies used block randomization and three studies used simple randomization. |
| ^5^ | Allocation concealment was adequate in all trials. All studies described the baseline characteristics of participants in all intervention groups. No serious baseline imbalance was observed. |
| ^6^ | All studies reported the number of loss to follow-up in all intervention groups. No serious differential loss to follow-up occurred for whole clusters or individuals in a cluster. |
| ^7^ | All studies adjusted for clustering in their analysis. |
| ^8^ | Moderate heterogeneity was observed in the pooled analysis (*I^2^* < 50%). |
| ^9^ | Laboratory-confirmed influenza was the outcome for all studies. |
| ^10^ | Confidence intervals are around estimates of effect that include both appreciable benefit and appreciable harm. |

### **Table S6. Grade quality assessment and evidence profile for face masks**

| **Quality assessment** | | | | | | | | **No. of patients** | | **Effect** |  |  |
| --- | --- | --- | --- | --- | --- | --- | --- | --- | --- | --- | --- | --- |
| No. of studies | | Design | Risk of bias | Inconsistency | Indirectness | Imprecision | Other considerations | Face mask | Control | Risk ratio | Quality | Importance |
| **Effect of hand hygiene intervention on prevention of laboratory-confirmed influenza** | | | | | | | |  |  |  |  |  |
| 7 | | Randomized  controlled trial^1,2^ | No serious risk of bias^3-7^ | Not serious^8^ | No serious indirectness^9^ | Serious^10^ | None | 130/2080 | 142/2167 | 0.94  (0.75, 1.17) | Moderate | Important |
| ^1^ | All studies were randomized controlled trials. | | | | | | | | | | | |
| ^2^ | All studies were cluster-RCTs | | | | | | | | | | | |
| ^3^ | Six studies reported blinding of study staff including clinical staff, laboratory staff or recruiting physicians. Subjects of all studies were not blinded due to the nature of the study design. | | | | | | | | | | | |
| ^4^ | Two studies used block randomization and five studies used simple randomization. | | | | | | | | | | | |
| ^5^ | Allocation concealment was adequate in all trials. All studies described the baseline characteristics of participants in all intervention groups. No serious baseline imbalance was observed. | | | | | | | | | | | |
| ^6^ | All studies reported the number of loss to follow-up in all intervention groups. No serious differential loss to follow-up occurred for whole clusters or individuals in a cluster. | | | | | | | | | | | |
| ^7^ | All studies adjusted for clustering in their analysis. | | | | | | | | | | | |
| ^8^ | Low heterogeneity was observed in the pooled analysis (*I^2^* < 30%). | | | | | | | | | | | |
| ^9^ | Laboratory-confirmed influenza was the outcome for all studies. | | | | | | | | | | | |
| ^10^ | Six confidence intervals are around estimates of effect that include both appreciable benefit and appreciable harm. | | | | | | | | | | | |
